# Effectiveness of COVID-19 Vaccines and Post-vaccination SARS-COV 2 Infection, Hospitalization, and Mortality: a Systematic Review and Meta-analysis of Observational Studies

**DOI:** 10.1101/2021.11.03.21265819

**Authors:** Kazem Rahmani, Rasoul Shavaleh, Mahtab Forouhi, Hamideh Feiz Disfani, Mostafa Kamandi, Aram Asareh Zadegan Dezfuli, Rozita Khatamian Oskooi, Molood Foogerdi, Moslem Soltani

## Abstract

**Introduction & Objective:** Vaccination is one of the most important and effective ways of preventing infectious diseases, and has recently been used in the COVID-19 epidemic and pandemic. The present meta-analysis study aimed to evaluate the effectiveness of COVID-19 vaccines in reducing the incidence of infection, hospitalization, and mortality in observational studies.

**Materials and Methods:** A systematic search was performed independently in Scopus, PubMed, ProQuest, and Google Scholar electronic databases as well as Preprint servers using the keywords under study. The heterogeneity of the studies was assessed using *I*^*2*^ *and* χ^*2*^ statistics, according to which the I^2^ of > 50% and P -value <0.1 was reported as heterogeneity of the studies. In addition, the Pooled Vaccine Effectiveness (PVE) obtained from the studies was calculated by converting (1-Pooled estimate × 100%) based on the type of outcome.

**Results:** A total of 54 records were included in this meta-analysis. The rate of PVE against SARS-COV 2 infection was about 71% (OR = 0.29, 95% CI: 0.23-0.36) in the first dose and 87% (OR = 0.13, 95% CI: 0.08-0.21) in the second, and the highest effectiveness in the first and second doses was that of BNT162b2 mRNA and combined studies. The PVE versus COVID-19-associated hospitalization was 73% (OR = 0.27, 95% CI: 0.18-0.41) in the first dose and 89% (OR = 0.11, 95% CI: 0.07-0.17) in the second. mRNA-1273 and combined studies in the first dose and ChAdOx1 and mRNA-1273 in the second dose had the highest effectiveness. Regarding the COVID-19-related mortality, PVE was about 28% (HR = 0.39, 95% CI: 0.23-0.45) in the first dose and 89% (HR = 0.11, 95% CI: 0.03-0.43) in the second.

**Conclusion:** The evidence obtained from this study showed that the effectiveness of BNT162b2 mRNA, mRNA-1273, and ChAdOx1 in the first and second doses, and even combined studies were associated with increased effectiveness against SARS-COV2 infection, hospitalization, and death from COVID-19. In addition, considering that the second dose was significantly more efficient than the first one, a booster dose injection could be effective in high-risk individuals. On the other hand, it was important to observe other prevention considerations in the first days after taking the first dose.

## Introduction

Over the past years, emerging and re-emerging diseases were a public health challenge in human populations with high mortality rates (1). In December 2019, a new outbreak of coronavirus by SARS coronavirus 2 (SARS-CoV-2) was reported in Wuhan, China, and on March 11, 2020, the World Health Organization and the CDC (Center for Disease Control and Prevention) introduced it as COVID-19 (2-4). The disease has been diagnosed with more than 195 million cases worldwide and more than 4 million deaths. As a result of this pandemic, restrictions and programs to prevent and control the infection were implemented by governments around the world (5, 6). The rapid spread of infection among individuals, the lack of symptoms or mild ones during the incubation period, and even the contagious nature of the disease during incubation have made the epidemic extremely difficult to control. Hence, most prevention programs were concentrated on making vaccine against SARS -CoV-2, and in several countries, some vaccines were licensed for emergency use (7-9). Clinical trials of manufactured vaccines showed that the effectiveness of Oxford-AstraZeneca (ChAdOx1), Pfizer BioNTech (BNT162b2 mRNA), Moderna (mRNA-1273), and Johnson & Johnson (Ad26.COV2.S) vaccines was 70.4%, 95%, 94.1%, and 66.9%, respectively (10-13). On the other hand, it should be noted that clinical trials are designed under controlled conditions for the voluntary entry of certain individuals and groups and the availability of the data of censored individuals (14), which can differ from the general population (15, 16). Furthermore, several observational studies were designed and conducted to determine the effectiveness of COVID-19 vaccines in various populations and groups under mass vaccination in order to not only indicate the effectiveness of COVID-19 vaccines, but also to compare the incidence of infection, mortality, and hospitalization due to COVID-19 in larger sample sizes and with more follow-ups (17-19). In the meantime, considering the wide range of evidence obtained from the effectiveness of vaccination in different groups, it seemed necessary to summarize them through meta-analysis studies to be presented to policy makers. Thus, the aim of this study was to evaluate the effectiveness of COVID-19 vaccines, the incidence of SARS-COV-2 infection, and the hospitalization and COVID-19-related mortality rates after vaccination in observational studies.

## Materials and methods

### Search Strategy

A systematic search of PubMed, Scopus, ProQuest, and Google Scholar databases as well as the Preprint servers including medRxiv and Research Square was done to identify the studies related to the keywords studied based on the Medical Subject Headings (MeSH) published until October 15, 2021, with full texts in English, without any spacial restrictions. The search was done blindly and independently by two researchers (K Rahmani & R Shavale) using the following keywords in the abovementioned databases: COVID-19; SARS-COV-2; coronavirus; vaccine; post-vaccination; mortality; hospitalization; readmission; reinfection; morbidity; and Breakthrough infections. The non-common cases that differed in the searches of the two researchers were dealt with by other researchers. Duplicates were also identified by title, author’s name, and journal name.

### Eligibility Criteria

All of the observational and longitudinal studies published in English that had assessed the effectiveness, incidence of SARS-COV 2 infection, hospitalization rates, and mortality rates after COVID-19 vaccination were included without any special restrictions. The studies that had examined the confirmed cases of SARS-COV 2 infection from positive real-time Reverse Transcription Polymerase Chain Reaction (RT-PCR or PCR) tests were also included, and the antibody-based studies and the ones based on other diagnostic methods were excluded from the review process. In addition, Case report, Case series, Letter or Correspondence, Animal studies, and studies with Mathematical model analysis were also excluded (flowchart 1). The studies on autoimmune, immunosuppressed, dialysis patients, or the patients with kidney problems and mental disorders in whom the severity of the disease varied were excluded as well. The studies that lacked an unvaccinated group to compare the results were not included in the review process. The studies on inactive vaccines such as CoronaVac and Covaxin as well as Ad26.COV2.S were excluded from the analysis due to their low frequencies.

### Outcomes

The outcomes were as follows:

1. Effectiveness of the vaccines against infection in the subjects studied, which was a relative reduction of RT-PCR test confirmed by throat swab, nasal swab, oropharyngeal swab, or saliva and sputum for Covid-19 in the vaccinated groups compared to the unvaccinated ones.
2. Effectiveness of the vaccines against hospitalization of the subjects in the studies was a relative reduction of hospitalization of the individuals whose RT-PCR test 14 days before and after hospitalization was confirmed through throat swab, nasal samples, oropharyngeal swab, or saliva and sputum for Covid-19 disease in the vaccinated groups compared with the unvaccinated ones.
3. Effectiveness of the vaccines against the hospitalization of the subjects in the studies was a relative reduction in deaths within 40 days after the RT-PCR test confirmed by throat swab, nasal swab, oropharyngeal swab, and or saliva and sputum for Covid-19 disease in the vaccinated groups compared with the unvaccinated ones (20).

### Data extraction

The data of the studies presented in flowchart 1 were extracted separately by two researchers. The extracted information included the names of the corresponding authors, types of vaccines studied, places of study, types of study, description of study conditions including study groups, positive SARS-COV 2 test cases in vaccinated and unvaccinated groups by the first and second doses, cases of death and hospitalization associated with COVID-19 in vaccinated and unvaccinated groups by the first and second doses, and infection and death HRs due to COVID-19 with 95% confidence interval between the first and second doses.

The HR of the studies was considered as the risk ratio of the vaccinated to non-vaccinated individuals, and in the studies whose HR was calculated as the risk ratio of unvaccinated to vaccinated people, it was converted with 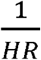 and 95% confidence intervals. Also, in the studies that had mentioned the effectiveness percentage through 1-HR × 100%, the HR and 95% of confidence intervals were converted by calculating 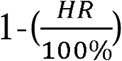. On the other hand, the follow-up periods in the studies were considered based on Pearson-Day, even in the studies where the follow-up periods were Pearson-Week and Pearson-Year.

Considering the studies examined, the people who had not taken any vaccines were classified as unvaccinated, and those who were on the ≥ 7^th^ day after the first dose and ≥ 5^th^ day after the second dose were classified as partial vaccinated and fully vaccinated, respectively.

### Statistical Analysis

A meta-analysis was carried out using the Random-effects Model and the Mantel-Haenszel weighting method for each study to estimate Pooled Odds Ratio, Pooled Hazard Ratio, and Pooled Incidence Ratio, and 95% confidence interval was used for the studies with similar effect measures (OR, IRR or HR). The heterogeneity of the studies was assessed using the *I*^*2*^ *and* χ^*2*^ statistics, according to the results of which *I*^*2*^ *>* 50% with P -value <0.1 showed the heterogeneity of the studies. In addition, to calculate the Pooled Vaccine Effectiveness (PVE) obtained from the studies, the following conversions were used: 1-Pooled Odds Ratio × 100%, 1-Pooled Hazard Ratio × 100%, and 1-Pooled Rate Ratio × 100%. The statistical analysis was performed using R version 4.1.1 and Metafor Package.

## Results

Based on the research strategy, 54 records on the effectiveness, incidence of SARS-COV 2 infection, mortality, and hospitalization associated with COVID-19 vaccination were finally reviewed (flowchart 1). The studies were analyzed based on the type of indices extracted. BNT162b2 mRNA accounted for the most frequent studies on vaccine types. In terms of location, most of the studies had been conducted in the USA, UK, Spain, and Israel, respectively (Tables 1 to 5). All of the studies had been carried out on the age groups >14 years.

**Table 1:** Incidence rate of SARS-COV 2 infection after the first and second doses in people with a history of COVID-19 vaccination

| First Author | Country | type of vaccines | type of study | Group of study | SARS-COV 2 Incidence |  |  |  |  |  |  |  |  |  |
| --- | --- | --- | --- | --- | --- | --- | --- | --- | --- | --- | --- | --- | --- | --- |
|  |  |  |  |  | Partial vaccinated |  | Unvaccinated |  | Rate Ratio* | Full Vaccinated |  | Unvaccinated |  | Rate Ratio* |
|  |  |  |  |  | Cases | Pearson day | Cases | Pearson day |  | Cases | Pearson day | Cases | Pearson day |  |
| Noa Dagan (23) | Israel | BNT162b2 mRNA | prospective cohort study | age of $\geq 16$ and older | 3533 | 262180 | 3971 | 261625 | 0.888 | - | - | - | - | - |
| Hall V FFPH (29) | UK | BNT162b2 mRNA | prospective cohort study | Healthcare workers. | 71 | 87278 | 977 | 710587 | 0.592 | 9 | 20978 | 977 | 710587 | 0.312 |
| Massimo Fabiani (30) | Italy | BNT162b2 mRNA | retrospective cohort | Healthcare workers. | 60 | 73914 | 128 | 62331 | 0.395 | 0 | 35596 | 11 | 14186 | 0.000 |
| Eric J Haas (17) | Israel | BNT162b2 mRNA | prospective cohort study | Age of 16 and older. | - | - | - | - |  | 6266 | 201882183 | 109876 | 120076136 | 0.034 |
| M.G. Thompson (31) | USA | BNT162b2 mRNA | prospective cohort study | health care workers | 8 | 49516 | 156 | 127971 | 0.133 | 3 | 120653 | 156 | 127971 | 0.020 |
| Sara Y Tartof (18) | USA | BNT162b2 mRNA | retrospective cohort | Health care workers. | 585 | 12986040 | 160280 | 449593130 | 0.126 | 3414 | 109442695 | 160280 | 449593130 | 0.088 |
| Madhumita Shrotri (32) | UK | BNT162b2 mRNA | prospective cohort study | aged 65 years and older | 50 | 17690 | 723 | 338003 | 1.321 | - | - | - | - | - |
| Yoel Angel (9) | Israel | BNT162b2 mRNA | retrospective cohort | healthcare workers | 68 | 117389 | 45 | 15091 | 0.194 | 27 | 168571 | 55 | 25359 | 0.074 |
| Colin Pawlowski (33) | USA | BNT162b2 mRNA | retrospective cohort | Aged $\geq 18$ years. | 401 | 2942986 | 1232 | 2851069 | 0.315 | 82 | 1914500 | 563 | 1828464 | 0.139 |
| Gili Regev-Yochay (34) | Israel | BNT162b2 mRNA | prospective cohort study | healthcare workers | 30 | 54832 | 115 | 199126 | 0.947 | 19 | 329071 | 115 | 199126 | 0.100 |
| Arjun Puranik (35) | USA | BNT162b2 mRNA | retrospective cohort | aged $\geq 18$ years | 58 | 180675 | 69 | 180614 | 0.840 | 72 | 2332005 | 321 | 2526895 | 0.243 |
| Mark A. Katz (36) | Israel | BNT162b2 mRNA | prospective cohort study | healthcare workers | - | - | - | - |  | 4 | 68574 | 9 | 10027 | 0.065 |
| Carmen Cabezas (37) | Catalonia | BNT162b2 mRNA | prospective cohort study | healthcare workers | 358 | 5528745 | 1961 | 2269003 | 0.075 | 222 | 4877162 | 961 | 2269003 | 0.107 |
| Aharona Freedman (38) | Israel | BNT162b2 mRNA | retrospective cohort | Aged $\geq 16$ years | 7166 | 14289253 | 133994 | 119701675 | 0.448 | 1639 | 14060250 | 95655 | 89535711 | 0.109 |
| Galia Zacay (39) | Israel | BNT162b2 mRNA | retrospective cohort | aged $\geq 16$ years | 59 | 28727 | 382 | 71797 | 0.386 | 15 | 26260 | 382 | 71797 | 0.107 |
| Victoria Jane Hall (40) | UK | BNT162b2 mRNA | prospective cohort study | healthcare workers | 71 | 87278 | 977 | 710587 | 0.592 | 9 | 20978 | 977 | 710587 | 0.312 |
| Hanne-Dorthe Emborg (41) | Denmark | BNT162b2 mRNA | prospective cohort study | prioritized risk groups, | 610 | 4415441 | 13297 | 55131206 | 0.573 | 304 | 13612638 | 13297 | 55131206 | 0.093 |
| Jonas Björk (42) | Sweden | BNT162b2 mRNA | prospective cohort study | aged 18-64 years, | 25 | 102830 | 4155 | 9902620 | 0.579 | 8 | 133616 | 4155 | 9902620 | 0.143 |
| Susana Monge (43) | Spain | BNT162b2 mRNA | prospective cohort study | aged $\geq 65$ years | 2690 | 218621 | 22 | 2128 | 1.190 | 885 | 207774 | 20 | 1997 | 0.425 |
| Madhumita Shrotri (32) | UK | ChAdOx1 | prospective cohort study | aged $\geq 65$ years | 82 | 32672 | 723 | 338003 | 1.173 | - | - | - | - | - |
| Subhadeep Ghosh (44) | India | ChAdOx1 | prospective cohort study | healthcare workers | 1159 | 49653918 | 10061 | 106594492 | 0.247 | 2512 | 58674639 | 10061 | 106594492 | 0.454 |
| M.G. Thompson (31) | USA | mRNA-1273 | prospective cohort study | health care workers | 3 | 31231 | 156 | 127971 | 0.079 | 2 | 40394 | 156 | 127971 | 0.041 |
| Colin Pawlowski (33) | USA | mRNA-1273 | retrospective cohort | Aged $\geq 18$ years. | 97 | 946890 | 303 | 927716 | 0.314 | 7 | 495550 | 101 | 478322 | 0.067 |
| Arjun Puranik (35) | USA | mRNA-1273 | retrospective cohort | aged $\geq 18$ years | 74 | 180810 | 69 | 180614 | 1.071 | 38 | 2214873 | 321 | 2526895 | 0.135 |
| Mark G. Thompson (45) | USA | Combination† | prospective cohort study | healthcare workers | 8 | 41856 | 161 | 116657 | 0.138 | 3 | 78902 | 161 | 116657 | 0.028 |
| Ashley Fowlkes (46) | USA | Combination† | prospective cohort study | healthcare workers | - | - | - | - |  | 34 | 454832 | 194 | 181357 | 0.070 |
| Tara C. Bouton (47) | USA | Combination† | prospective cohort study | healthcare workers, | 29 | 251790 | 329 | 406387 | 0.142 | - | - | - | - | - |

\* Rate Ratio computed
† BNT162b2 mRNA and mRNA-1273 and ChAdOx1

**Table 2.** Positive tests for SARS-COV 2 infection after the first and second doses in people with a history of COVID-19 vaccination

| First Author | Country | type of vaccines | type of study | Group of study | Positive SARS-COV 2 Infection |  |  |  |  |  |  |  |  |  |
| --- | --- | --- | --- | --- | --- | --- | --- | --- | --- | --- | --- | --- | --- | --- |
|  |  |  |  |  | Partial vaccinated |  | Unvaccinated |  | Odds Ratio* | Full Vaccinated |  | Unvaccinated |  | Odds Ratio* |
|  |  |  |  |  | Cases | Total | Cases | Total |  | Cases | Total | Cases | Total |  |
| Hall V FFPH (29) | UK | BNT162b2 mRNA | prospective cohort study | Healthcare workers. | 71 | 20641 | 977 | 2683 | 0.006 | 9 | 1607 | 977 | 2683 | 0.009 |
| Iván Martínez-Baz (48) | Spain | BNT162b2 mRNA | prospective cohort study | aged ≥ 18 years | 90 | 310 | 6980 | 19580 | 0.738 | 61 | 491 | 6980 | 19580 | 0.256 |
| T. Pilishvili (49) | USA | BNT162b2 mRNA | case–control study | healthcare workers | 122 | 1472 | 707 | 3420 | 0.347 | 149 | 1472 | 882 | 3420 | 0.324 |
| Ping Ye, DNP (50) | USA | BNT162b2 mRNA | retrospective cohort | nursing home residents | 68 | 86 | 5 | 5 | 0.000 | 5 | 17 | 5 | 5 | 0.000 |
| Tamara Pilishvili (51) | USA | BNT162b2 mRNA | case–control study | health care workers | 214 | 926 | 340 | 642 | 0.267 | - | - | - | - | - |
| Iván Martínez-Baz (52) | Spain | BNT162b2 mRNA | prospective cohort study | aged ≥ 18 year | 351 | 2022 | 4811 | 14348 | 0.416 | 1070 | 7972 | 4811 | 14348 | 0.307 |
| Sara Carazo (53) | Canada | BNT162b2 mRNA | case–control study | healthcare workers | 2130 | 26719 | 6323 | 24986 | 0.256 | 68 | 2022 | 6323 | 24986 | 0.103 |
| Carmen Cabezas (37) | Catalonia | BNT162b2 mRNA | prospective cohort study | healthcare workers | 1607 | 102161 | 4440 | 116417 | 0.403 | - | - | - | - | - |
| Galia Zacay (39) | Israel | BNT162b2 mRNA | retrospective cohort | aged ≥16 years | 59 | 1445 | 382 | 6286 | 0.658 | 16 | 2941 | 382 | 6286 | 0.085 |
| Tariq Azamgarhi (54) | UK | BNT162b2 mRNA | prospective cohort study | healthcare workers | 23 | 1408 | 26 | 825 | 0.510 | - | - | - | - | - |
| Jamie Lopez Bernal (28) | UK | BNT162b2 mRNA | case–control study | age ≥70 years, | 448 | 1956 | 15287 | 36668 | 0.416 | - | - | - | - | - |
| Jamie Lopez Bernal (26) | UK | BNT162b2 mRNA | case–control study | aged ≥16 years, | 43 | 2884 | 4043 | 96371 | 0.346 | 122 | 15749 | 4043 | 96371 | 0.178 |
| T. Pilishvili (49) | USA | mRNA-1273 | case–control study | health care workers | 18 | 1472 | 156 | 3420 | 0.259 | 18 | 1472 | 190 | 3420 | 0.210 |
| Tamara Pilishvili (51) | USA | mRNA-1273 | case–control study | health care workers | 68 | 268 | 340 | 642 | 0.302 | - | - | - | - |  |
| Iván Martínez-Baz (52) | Spain | mRNA-1273 | prospective cohort | aged ≥ 18 year | 70 | 517 | 4811 | 14348 | 0.310 | 85 | 1127 | 4811 | 14348 | 0.162 |
| Sara Carazo (53) | Canada | mRNA-1273 | case–control study | healthcare workers | 110 | 1639 | 6323 | 24986 | 0.212 | 2 | 128 | 6323 | 24986 | 0.047 |
| Jamie Lopez Bernal (26) | UK | mRNA-1273 | case control | aged ≥16 years, | 592 | 25913 | 4043 | 96371 | 0.534 | 218 | 8244 | 4043 | 96371 | 0.620 |
| Saurabh Bobdey (55) | India | ChAdOx1 | prospective cohort study | - | 27 | 239 | 19 | 94 | 0.503 | 67 | 2863 | 19 | 94 | 0.095 |
| Iván Martínez-Baz (48) | Spain | ChAdOx1 | prospective cohort study | aged ≥ 18 years | 99 | 524 | 6980 | 19580 | 0.420 | - | - | - | - | - |
| Iván Martínez-Baz (52) | Spain | ChAdOx1 | prospective cohort study | aged ≥ 18 year - | 302 | 1599 | 4811 | 14348 | 0.462 | 272 | 1539 | 4811 | 14348 | 0.426 |
| Aleena Issac (56) | India | ChAdOx1 | prospective cohort study | Healthcare Workers | - | - | - | - | - | 16 | 243 | 35 | 80 | 0.091 |
| Jamie Lopez Bernal (28) | UK | ChAdOx1 | case–control study | adult age ≥70 years, | 396 | 1342 | 15287 | 36668 | 0.585 | - | - | - | - | - |
| Eli S. Rosenberg (57) | USA | Combination† | prospective cohort | adults aged ≥18 | - | - | - | - | - | 9675 | 10135322 | 38505 | 3742197 | 0.092 |
| Maria Elena Flacco (58) | Italy | Combination† | retrospective cohort | Aged ≥ 18 years. | 12 | 69539 | 6948 | 175687 | 0.004 | - | - | - | - | - |
| Aaron J. Tande (59) | USA | Combination† | retrospective cohort | aged ≥18 years | 42 | 3006 | 1436 | 45327 | 0.433 | - | - | - | - | - |
| Anoop S V Shah (60) | Scotland | Combination† | prospective cohort study | healthcare workers | 1152 | 109074 | 3191 | 144525 | 0.473 | - | - | - | - | - |
| Kristin L. Andrejko (61) | USA | Combination† | case–control study | aged ≥18 years | 51 | 150 | 454 | 767 | 0.355 | 20 | 106 | 454 | 767 | 0.160 |
| Nathanael Fillmore (62) | USA | Combination† | retrospective cohort | - | - | - | - | - | - | 1546 | 3627 | 6326 | 11569 | 0.616 |
| Tara C. Bouton (47) | USA | Combination† | prospective cohort study | healthcare workers | 96 | 7109 | 329 | 3481 | 0.131 | 17 | 5913 | 329 | 3481 | 0.028 |
| Hannah Chung (63) | Canada | Combination† | case–control study | aged ≥16 years | 2050 | 21272 | 51220 | 302761 | 0.524 | 73 | 21272 | 51220 | 302761 | 0.017 |
| Alyson Cavanaugh (64) | USA | Combination† | case–control study | aged ≥18 years | 17 | 56 | 179 | 463 | 0.692 | 50 | 219 | 179 | 463 | 0.469 |

\* Odds Ratio computed
† BNT162b2 mRNA and mRNA-1273 and ChAdOx1

**Table 3.** – Covid-19-related mortality after the first and second doses in people vaccinated with BNT162b2 mRNA

| First Author | Vaccine type | Country | type of study | Group of study | SARS-COV 2 Mortality |  |  |  |  |  |  |  |  |  |
| --- | --- | --- | --- | --- | --- | --- | --- | --- | --- | --- | --- | --- | --- | --- |
|  |  |  |  |  | Partial vaccinated |  | Unvaccinated |  | Rate Ratio* | Full Vaccinated |  | Unvaccinated |  | Rate Ratio* |
|  |  |  |  |  | death | Pearson day | death | Pearson day |  | death | Pearson day | death | Pearson day |  |
| Noa Dagan (23) | BNT162b2 mRNA | Israel | prospective cohort study | age of ≥ 16 and older | 2 | 264538 | 6 | 264479 | 0.333 | 9 | 4322 | 32 | 4316 | 0.281 |
| Eric J Haas (17) | BNT162b2 mRNA | Israel | prospective cohort study | Age of 16 and older. | - | - | - | - | - | 138 | 201882183 | 715 | 120076136 | 0.115 |
| Arjun Puranik (35) | BNT162b2 mRNA | USA | retrospective cohort | aged ≥18 years | 0 | 180814 | 0 | 180819 | - | 0 | 2333860 | 4 | 2537030 | 0.000 |
| Carmen Cabezas (37) | BNT162b2 mRNA | Catalonia | prospective cohort study | healthcare workers | 153 | 3030779 | 272 | 639181 | 0.119 | 33 | 2745713 | 272 | 639181 | 0.028 |
| Aharona Freedman (38) | BNT162b2 mRNA | Israel | retrospective cohort | Aged ≥16 years | 178 | 14289253 | 819 | 119701675 | 1.821 | 61 | 14060250 | 567 | 89535711 | 0.685 |
| Hanne-Dorthe Emborg (41) | BNT162b2 mRNA | Denmark | prospective cohort study | - | 203 | 4551842 | 445 | 55910554 | 5.603 | 25 | 13735570 | 445 | 55910554 | 0.229 |
| Arjun Puranik (35) | mRNA-1273 | USA | retrospective cohort | aged ≥18 years | 0 | 180951 | 0 | 180819 | - | 0 | 2215773 | 4 | 2537030 | 0.000 |
| Subhadeep Ghosh (44) | ChAdOx1 | India | prospective cohort study | healthcare workers | 16 | 49653918 | 37 | 106594492 | 0.928 | 7 | 58674639 | 37 | 106594492 | 0.344 |
| Baltazar Nunes (65) | Combination† | Portugal | prospective cohort study | aged ≥65 years | 11 | 21560915 | 90 | 52945805 | 0.300 | 14 | 48853060 | 90 | 52945805 | 0.169 |

\* Rate Ratio computed
† BNT162b2 mRNA and mRNA-1273 and ChAdOx1

**Table 4:**
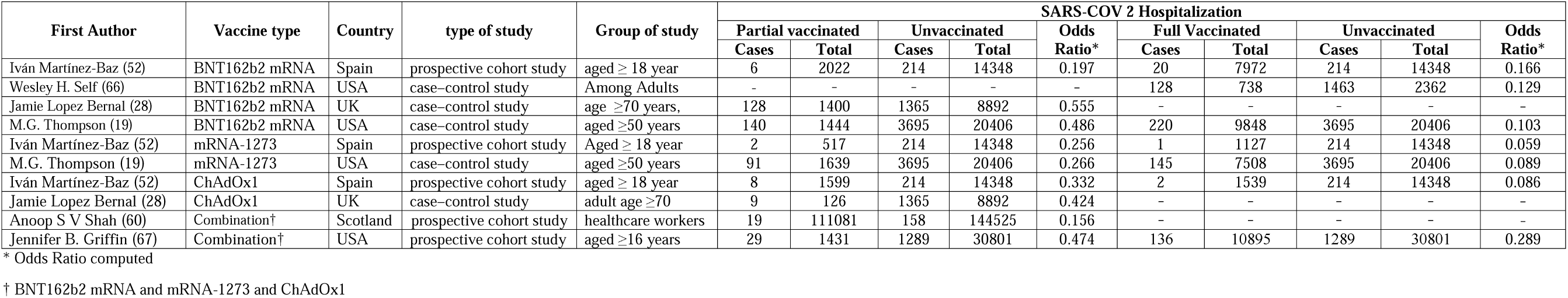
COVID-19-related hospitalization rate after the first and second doses of vaccinated patients

| First Author | Vaccine type | Country | type of study | Group of study | SARS-COV 2 Hospitalization |  |  |  |  |  |  |  |  |  |
| --- | --- | --- | --- | --- | --- | --- | --- | --- | --- | --- | --- | --- | --- | --- |
|  |  |  |  |  | Partial vaccinated |  | Unvaccinated |  | Odds Ratio* | Full Vaccinated |  | Unvaccinated |  | Odds Ratio* |
|  |  |  |  |  | Cases | Total | Cases | Total |  | Cases | Total | Cases | Total |  |
| Iván Martínez-Baz (52) | BNT162b2 mRNA | Spain | prospective cohort study | aged ≥ 18 year | 6 | 2022 | 214 | 14348 | 0.197 | 20 | 7972 | 214 | 14348 | 0.166 |
| Wesley H. Self (66) | BNT162b2 mRNA | USA | case–control study | Among Adults | - | - | - | - | - | 128 | 738 | 1463 | 2362 | 0.129 |
| Jamie Lopez Bernal (28) | BNT162b2 mRNA | UK | case–control study | age ≥70 years, | 128 | 1400 | 1365 | 8892 | 0.555 | - | - | - | - | - |
| M.G. Thompson (19) | BNT162b2 mRNA | USA | case–control study | aged ≥50 years | 140 | 1444 | 3695 | 20406 | 0.486 | 220 | 9848 | 3695 | 20406 | 0.103 |
| Iván Martínez-Baz (52) | mRNA-1273 | Spain | prospective cohort study | Aged ≥ 18 year | 2 | 517 | 214 | 14348 | 0.256 | 1 | 1127 | 214 | 14348 | 0.059 |
| M.G. Thompson (19) | mRNA-1273 | USA | case–control study | aged ≥50 years | 91 | 1639 | 3695 | 20406 | 0.266 | 145 | 7508 | 3695 | 20406 | 0.089 |
| Iván Martínez-Baz (52) | ChAdOx1 | Spain | prospective cohort study | aged ≥ 18 year | 8 | 1599 | 214 | 14348 | 0.332 | 2 | 1539 | 214 | 14348 | 0.086 |
| Jamie Lopez Bernal (28) | ChAdOx1 | UK | case–control study | adult age ≥70 | 9 | 126 | 1365 | 8892 | 0.424 | - | - | - | - | - |
| Anoop S V Shah (60) | Combination† | Scotland | prospective cohort study | healthcare workers | 19 | 111081 | 158 | 144525 | 0.156 | - | - | - | - | - |
| Jennifer B. Griffin (67) | Combination† | USA | prospective cohort study | aged ≥16 years | 29 | 1431 | 1289 | 30801 | 0.474 | 136 | 10895 | 1289 | 30801 | 0.289 |

\* Odds Ratio computed
† BNT162b2 mRNA and mRNA-1273 and ChAdOx1

**Table 5.** Risk ratio of SARS-COV 2 infection and COVID-19-related mortality in patients with a history of first- and second-dose vaccination

| First Author | Vaccine type | Country | type of study | Group of study | HR SARS-COV 2 infection |  |  |  |  |  | HR death related to the COVID-19 |  |  |  |  |  |
| --- | --- | --- | --- | --- | --- | --- | --- | --- | --- | --- | --- | --- | --- | --- | --- | --- |
|  |  |  |  |  | Partial vaccinated |  |  | Full Vaccinated |  |  | Partial vaccinated |  |  | Full Vaccinated |  |  |
|  |  |  |  |  | HR* | 95% CI |  | HR* | 95% CI |  | HR* | 95% CI |  | HR* | 95% CI |  |
| Hall V FFPH (29) | BNT162b2 mRNA | UK | prospective cohort study | Healthcare workers. | 0.3 | 0.15 | 0.45 | 0.15 | 0.04 | 0.26 | - | - | - | - | - | - |
| Amadea Britton (68) | BNT162b2 mRNA | USA | retrospective cohort | - | 0.37 | 0.21 | 0.67 | - | - | - | - | - | - | - | - | - |
| Adeel A. Butt (69) | BNT162b2 mRNA | Qatar | prospective cohort study | - | - | - | - | - | - | - | - | - | - | 0.35 | 0.22 | 0.55 |
| Ioannis Baltas (70) | BNT162b2 mRNA | UK | case–control study | - | - | - | - | - | - | - | 0.34 | 0.178 | 0.651 |  |  |  |
| M.G. Thompson (31) | BNT162b2 mRNA | USA | prospective cohort study | healthcare workers | 0.2 | 0.1 | 0.4 | 0.07 | 0.02 | 0.22 | - | - | - | - | - | - |
| Sara Y Tartof (18) | BNT162b2 mRNA | USA | retrospective cohort | Aged $\geq 12$ years. | 0.42 | 0.39 | 0.46 | 0.27 | 0.26 | 0.28 | - | - | - | - | - | - |
| Madhumita Shrotri (32) | BNT162b2 mRNA | UK | prospective cohort study | age of $\geq 65$ | 0.77 | 0.37 | 1.58 | - | - | - | - | - | - | - | - | - |
| Mark A. Katz (36) | BNT162b2 mRNA | Israel | prospective cohort study | healthcare workers | - | - | - | 0.055 | 0.018 | 0.174 | - | - | - | - | - | - |
| Jamie Lopez Bernal (71) | BNT162b2 mRNA | UK | retrospective cohort | aged $\geq 70$ years | - | - | - | - | - | - | 0.56 | 0.47 | 0.68 | 0.31 | 0.14 | 0.69 |
| Ben Glampson (72) | BNT162b2 mRNA | UK | retrospective cohort | Aged $\geq 16$ years. | 0.42 | 0.36 | 0.5 | - | - | - | - | - | - | - | - | - |
| Carmen Cabezas (37) | BNT162b2 mRNA | Catalonia | prospective cohort study | healthcare workers | 0.13 | 0.11 | 0.14 | 0.13 | 0.11 | 0.16 | 0.31 | 0.26 | 0.39 | 0.03 | 0.02 | 0.04 |
| Tariq Azamgarhi (54) | BNT162b2 mRNA | UK | prospective cohort study | healthcare workers | 0.3 | 0.09 | 0.94 | - | - | - | - | - | - | - | - | - |
| Jamie Lopez Bernal (28) | BNT162b2 mRNA | UK | case–control study | adult age $\geq 70$ years, | - | - | - | - | - | - | 0.49 | 0.38 | 0.63 | - | - | - |
| Ida Rask Moustsen-Helms (73) | BNT162b2 mRNA | Denmark | retrospective cohort | healthcare workers, | 0.17 | 0.04 | 0.28 | 0.9 | 0.82 | 0.95 | - | - | - | - | - | - |
| Peter Nordstrom (24) | BNT162b2 mRNA | Sweden | Prospective cohort | - | - | - | - | 0.22 | 0.21 | 0.22 | - | - | - | - | - | - |
| M.G. Thompson (31) | mRNA-1273 | USA | prospective cohort study | health care workers | 0.17 | 0.05 | 0.6 | 0.18 | 0.04 | 0.8 | - | - | - | - | - | - |
| Peter Nordstrom (24) | mRNA-1273 | Sweden | Prospective cohort | - | - | - | - | 0.13 | 0.12 | 0.16 | - | - | - | - | - | - |
| Saurabh Bobdey (55) | ChAdOx1 | India | prospective cohort study | - | 0.559 | 0.327 | 0.954 | 0.114 | 0.0763 | 0.184 | - | - | - | - | - | - |
| Ioannis Baltas (70) | ChAdOx1 | UK | case–control study | - | - | - | - | - | - | - | 0.216 | 0.067 | 0.696 | - | - | - |
| Madhumita Shrotri (32) | ChAdOx1 | UK | prospective cohort study | aged $\geq 65$ years | 0.95 | 0.5 | 1.84 | - | - | - | - | - | - | - | - | - |
| Jamie Lopez Bernal (71) | ChAdOx1 | UK | retrospective cohort | aged $\geq 70$ years | - | - | - | - | - | - | 0.45 | 0.34 | 0.59 | - | - | - |
| Ben Glampson (72) | ChAdOx1 | UK | retrospective cohort | Aged $\geq 16$ years. | 0.59 | 0.49 | 0.71 | - | - | - | - | - | - | - | - | - |
| Peter Nordstrom (24) | ChAdOx1 | Sweden | Prospective cohort | - | - | - | - | 0.5 | 0.42 | 0.59 | - | - | - | - | - | - |
| Mark G. Thompson (45) | Combination† | USA | prospective cohort study | healthcare workers | 0.2 | 0.1 | 0.41 | 0.1 | 0.03 | 0.32 | - | - | - | - | - | - |
| Ashley Fowlkes (46) | Combination† | USA | prospective cohort study | healthcare workers |  |  |  | 0.2 | 0.12 | 0.31 | - | - | - | - | - | - |
| Sarah E. Waldman (74) | Combination† | USA | retrospective cohort | age $\geq 18$ year old | 0.53 | 0.4 | 0.71 | 0.22 | 0.12 | 0.42 | - | - | - | - | - | - |
| Maria Elena Flacco (58) | Combination† | Italy | retrospective cohort | Aged $\geq 18$ years. | 0.05 | 0.04 | 0.06 | 0.02 | 0.01 | 0.03 | 0.03 | 0.01 | 0.08 | 0.02 | 0 | 0.12 |
| Baltazar Nunes (65) | Combination† | Portugal | prospective cohort study | Aged $\geq 65$ years. | - | - | - | - | - | - | 0.23 | 0.12 | 0.44 | 0.04 | 0.02 | 0.08 |

\* Hazard Ratio adjusted in each study
† BNT162b2 mRNA and mRNA-1273 and ChAdOx1

### Effectiveness of Vaccines against Covid-19 Infection in Partial Vaccinated Group

The results of 28 studies presented as forest plot using effect measure pooled OR in Figure 1 showed that the effectiveness of the first dose (Partial) of the studied vaccines against COVID-19 infection was 71% in total. This effectiveness varied according to the type of vaccine (*p* − *value*_*subgroup*_ < 0.05); i.e. the effectiveness of BNT162b2 mRNA vaccine against COVID-19 infection was 72% (Pooled OR = 0.28, 95% CI: 0.13-0.60), the effectiveness of mRNA-1273 vaccine was 69% (Pooled OR = 0.31, 95% CI: 0.22-0.44), and that of ChAdOx1 vaccine was 51% (Pooled OR = 0.49 95% CI: 0.41-0.59). Furthermore, the combined studies that had examined the vaccines (BNT162b2 mRNA, mRNA-1273, ChAdOx1) reported an approximate effectiveness of 79% (Pooled OR = 0.21, 95% CI: 0.06-0.76) (Figure 1).

**Figure 1-.**
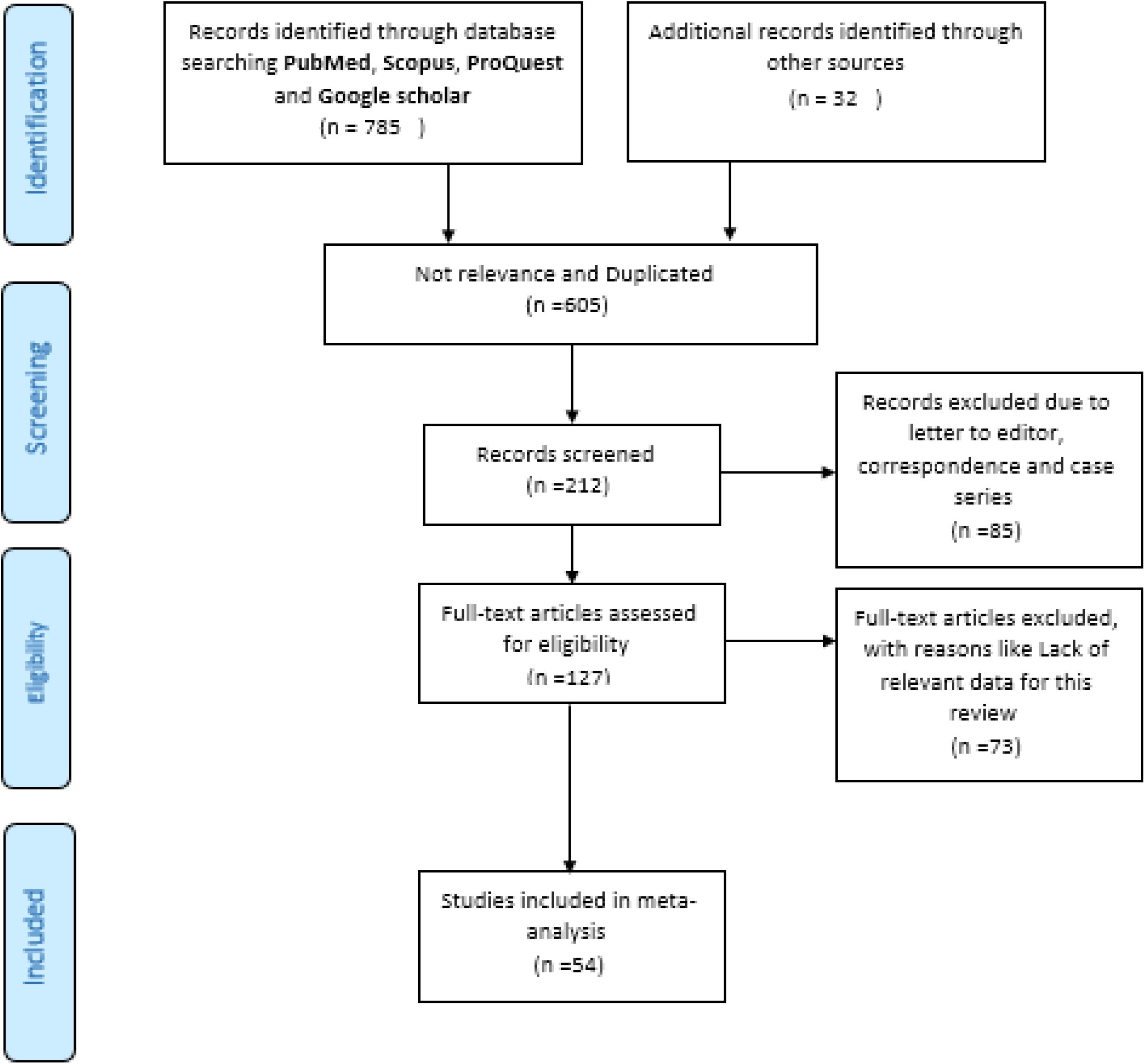
PRISMA flow diagram of the studies included in meta-analysis

**Figure 1.**
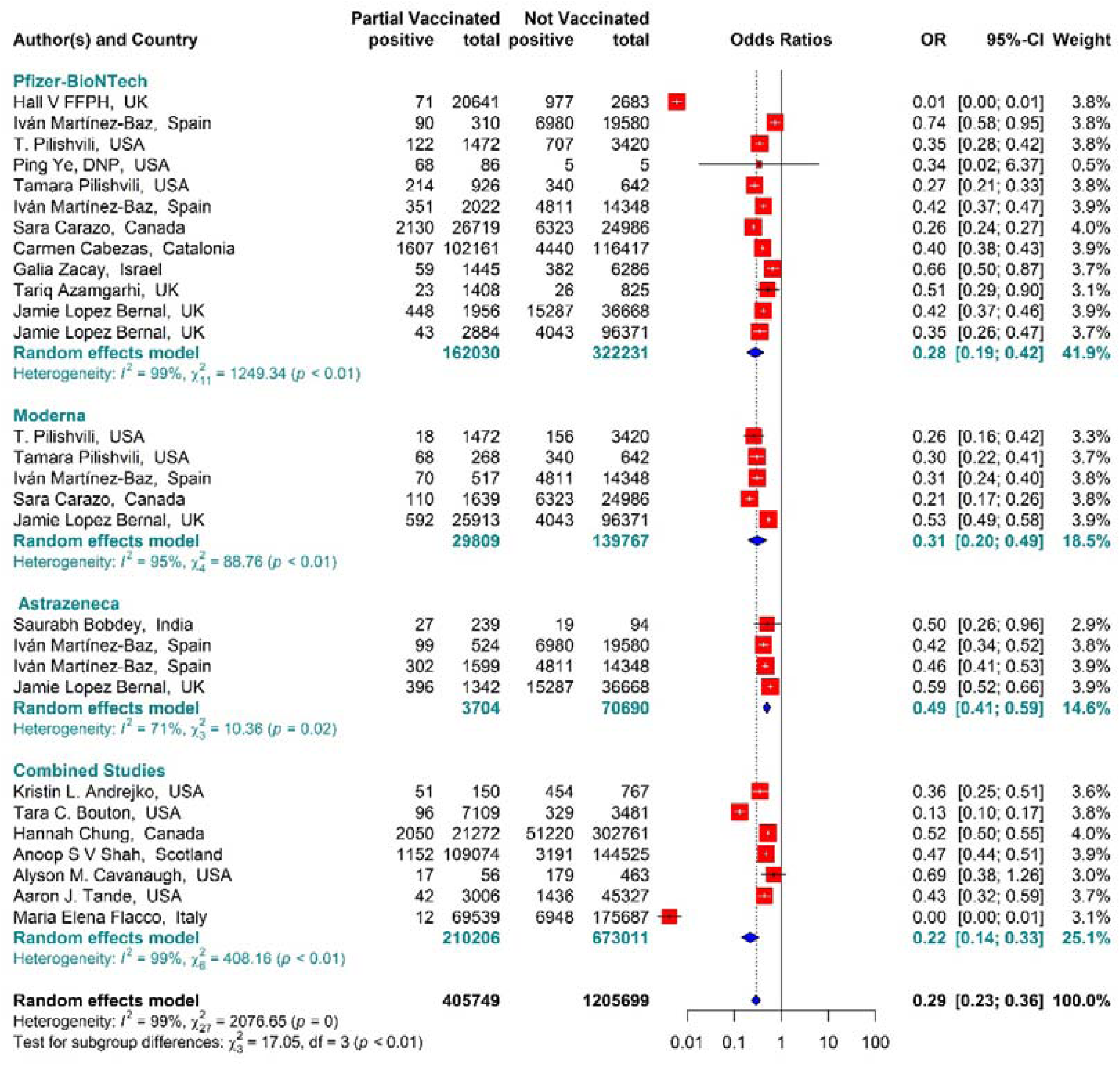
Partial vaccinated effectiveness of BNT162b2 mRNA, mRNA-1273, and ChAdOx1 vaccines against SARS-COV 2 infection

### Effectiveness of Vaccines against Covid-19 Infection in Full Vaccinated Group

The results of 20 studies presented as forest plot using effect measure pooled OR in Figure 2 showed that the total effectiveness of the second dose of the vaccines (Full vaccinated) against COVID-19 infection was 87% (OR = 0.13, 95% CI: 0.08-0.22); i.e. the effectiveness of the second dose of BNT162b2 mRNA vaccine against COVID-19 infection was 88% (OR = 0.12, 95% CI: 0.05-0.27), that of mRNA vaccine -1273 was 80% (OR = 0.20, 95% CI: 0.08-0.52), and the effectiveness of the second dose of ChAdOx1 vaccine was 5184% (OR = 0.16, 95% CI: 0.06 -0.45). In addition, the combined studies that examined the vaccines (BNT162b2 mRNA, mRNA-1273, ChAdOx1) reported an approximate effectiveness of 0.89% for the second doses (OR = 0.13, 95% CI: 0.08-0.22). The results of the sub group analysis in the second dose showed well that there was no difference between different types of vaccines in terms of their effectiveness (*p* − *value*_*subgroup*_ *=* 0.81) (Figure 2).

**Figure 2.**
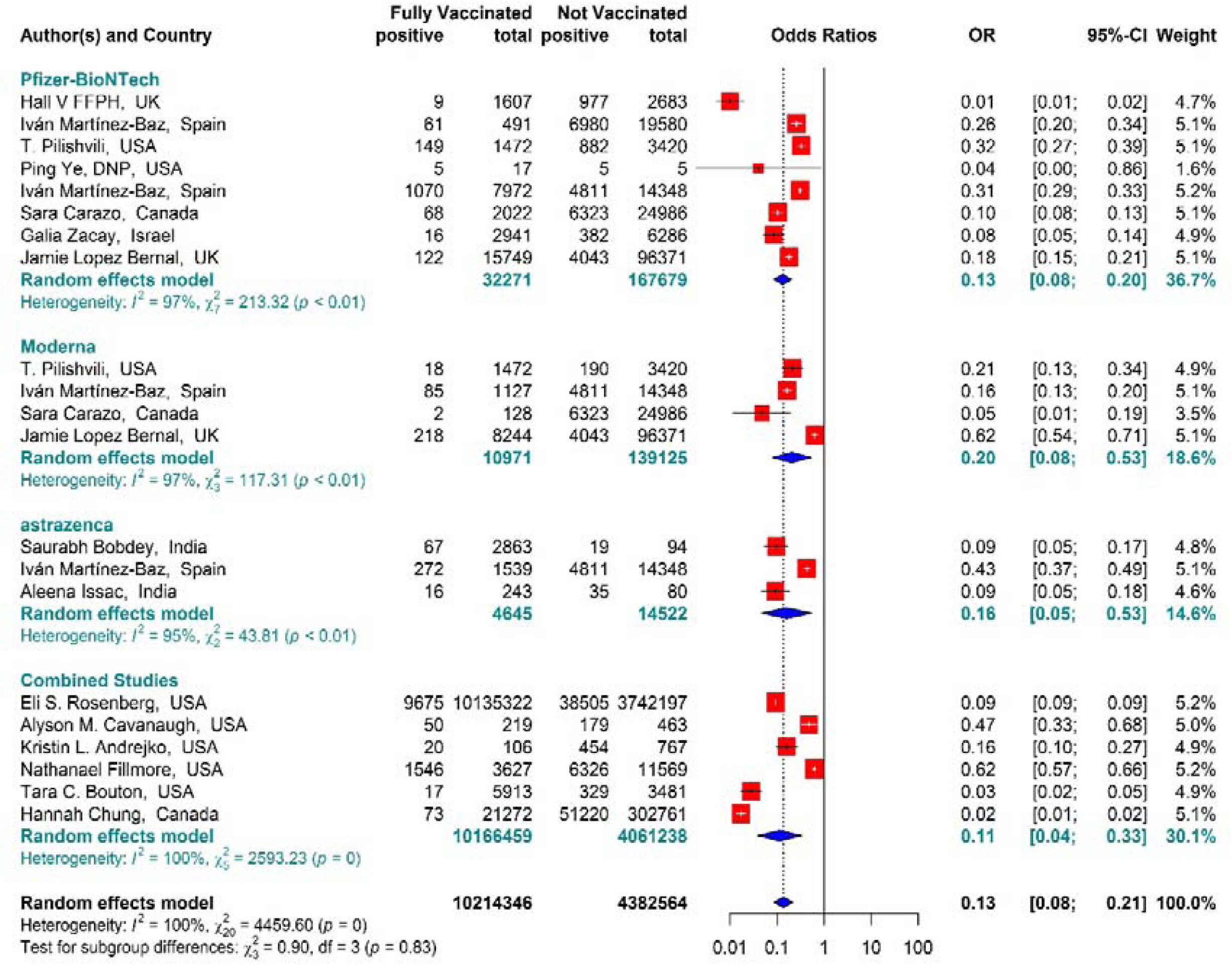
Full vaccinated effectiveness of BNT162b2 mRNA, mRNA-1273, and ChAdOx1 vaccines against SARS-COV 2 infection.

### Effectiveness of vaccines against hospitalization of COVID-19 patients in the Partial vaccinated group

The total effectiveness of BNT162b2 mRNA, mRNA-1273, and ChAdOx1 vaccines as well as the Combined studies in the first dose against COVID-19-related hospitalization was 73% (OR = 0.27, 95% CI: 0.17-0.44) (Figure 3). Considering the type of vaccines, the results of Pooled estimate showed that the effectiveness of BNT162b2 mRNA vaccine was about 85% (OR = 0.15, 95% CI: 0.04-0.56), that of mRNA-1273 was 73% (OR = 0.27, 95% CI: 0.21 - 0.34), and the effectiveness of ChAdOx1 vaccine was about 56% (OR = 0.44, 95% CI: 0.29-0.67). In the Combined studies, the efficacy of the vaccines was about 62% (OR = 0.38, 95% CI: 0.23-0.62). The results of the subgroup on the type of vaccines also indicated no difference between the effectiveness of the vaccines in the first dose against hospitalization with COVID-19 (P value = 0.11).

**Figure 3-.**
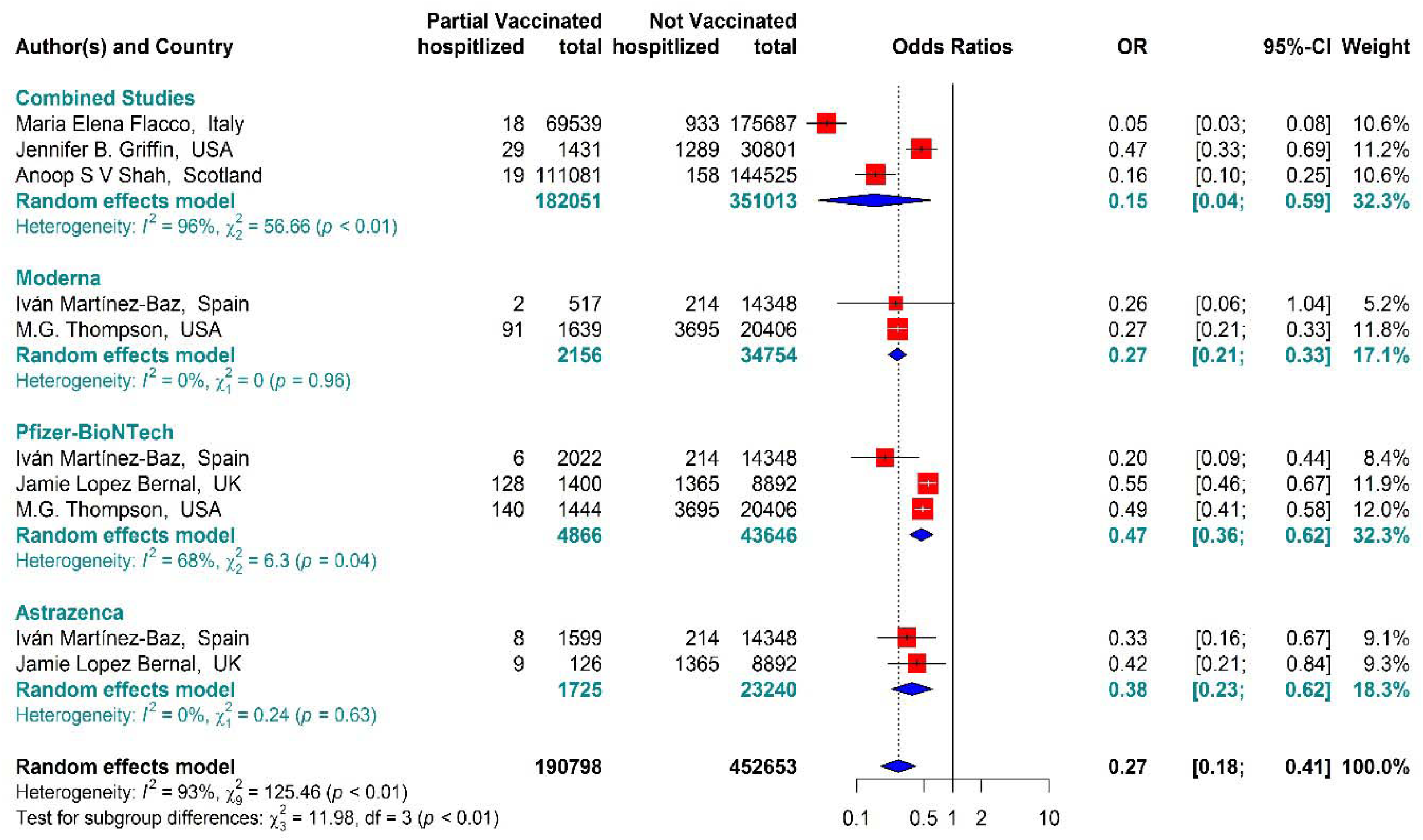
Partial effectiveness of BNT162b2 mRNA, mRNA-1273, and ChAdOx1 vaccines against COVID-19-related hospitalization

### Effectiveness of vaccines against hospitalization of COVID-19 patients in Full vaccinated group

The total effectiveness of BNT162b2 mRNA, mRNA-1273, and ChAdOx1 vaccines as well as the Combined Studies in the second dose against COVID-19-related hospitalization was about 89% (OR = 0.11, 95% CI: 0.08-0.16) while BNT162b2 mRNA, MRNA-1273, and ChAdOx1 vaccines had the effectiveness of 91%, 88%, and 91%, respectively. In addition, the effectiveness of the vaccines in the combined studies was about 86% (OR = 0.14, 95% CI: 0.03-0.60). The results of the sub group analysis in the second dose showed well that there was no difference between the effectiveness rates of different types of vaccines against hospitalization (*p* − *value*_*subgroup*_ = 0.09) (Figure 4).

**Figure 4-.**
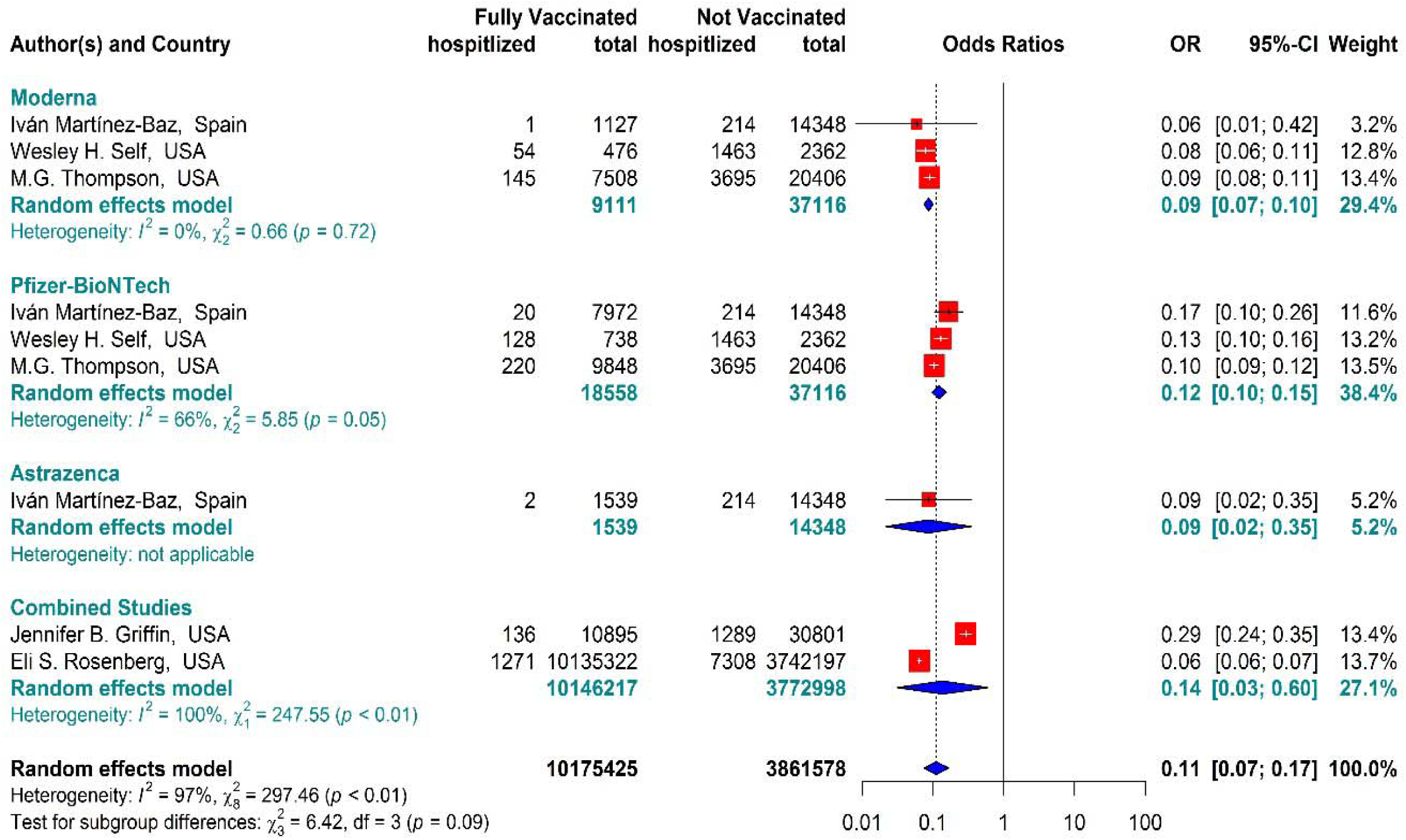
Full effectiveness of BNT162b2 mRNA, mRNA-1273, and ChAdOx1 vaccines against COVID-19-related hospitalization

### Effectiveness of vaccines against infection using COVID-19-associated IRR in partially vaccinated individuals

The results indicated that the rate of COVID-19-related infection in the people vaccinated with BNT162b2 mRNA, mRNA-1273, ChAdOx1, and Combined studies in the first dose was reduced by about 60% (IRR = 0.4, 95% CI: 0.30-0.53 (Figure 5). The reduction in COVID-19-related infection rate in the individuals vaccinated with the first dose of BNT162b2 mRNA, mRNA-1273, and ChAdOx1was about 56% (IRR = 0.44, 95% CI: 0.31-0.61), 66% (IRR = 0.34, 95% CI: 0.11-1.02), and 46% (IRR = 0.54, 95% CI: 0.12-2.48), respectively. In the Combined studies, the reduction in COVID-19-related infection was about 86% (IRR = 0.14, 95% CI: 0.10-0.20) as well. The results of the sub group analysis in the first dose showed well that there was a difference between the effectiveness rates of different types of vaccine against SARS-COV 2 infection (*p* − *value*_*subgroup*_ < 0.01) (Figure 5).

**Figure 5.**
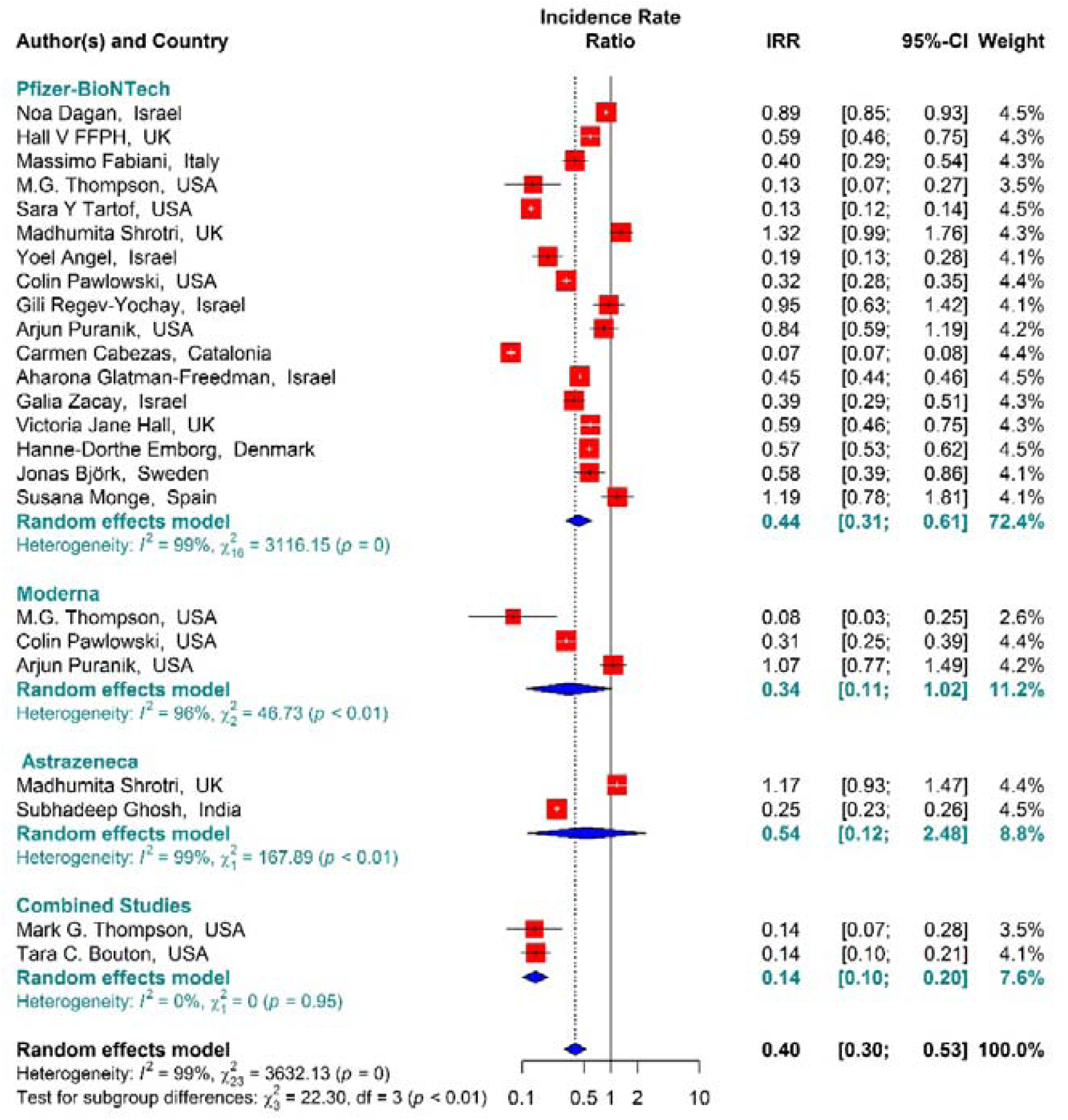
Effectiveness of vaccines against infection using COVID-19-related IRR in partial vaccinated individuals

### Effectiveness of vaccines using COVID-19-associated IRR in full-vaccinated individuals

In the people vaccinated with the second dose (Full vaccinated) of BNT162b2 mRNA, mRNA-1273, ChAdOx1, and Combined studies, the rate of COVID-19-related infection had reduced by about 90% (IRR = 0.10, 95% CI: 0.07-0.17) (Figure 6). The reduction in COVID-19-related infection rate in the individuals vaccinated with the second dose of BNT162b2 mRNA, mRNA-1273, and ChAdOx1was about 89% (IRR = 0.11, 95% CI: 0.08-0.16), 91% (IRR = 0.09, 95% CI: 0.04-0.17), and 55% (IRR = 0.45, 95% CI: 0.43-0.47), respectively (Figure 6). In the Combined studies, the COVID-19-related mortality after the second dose had reduced by about 95% (IRR = 0.05, 95% CI: 0.02-0.13). The results of the sub group analysis in the second dose showed well that there was a difference between the effectiveness rates of different types of vaccine against SARS-COV 2 infection (*p-value*_*subgroup*_ < 0.01) (Figure 6).

**Figure 6.**
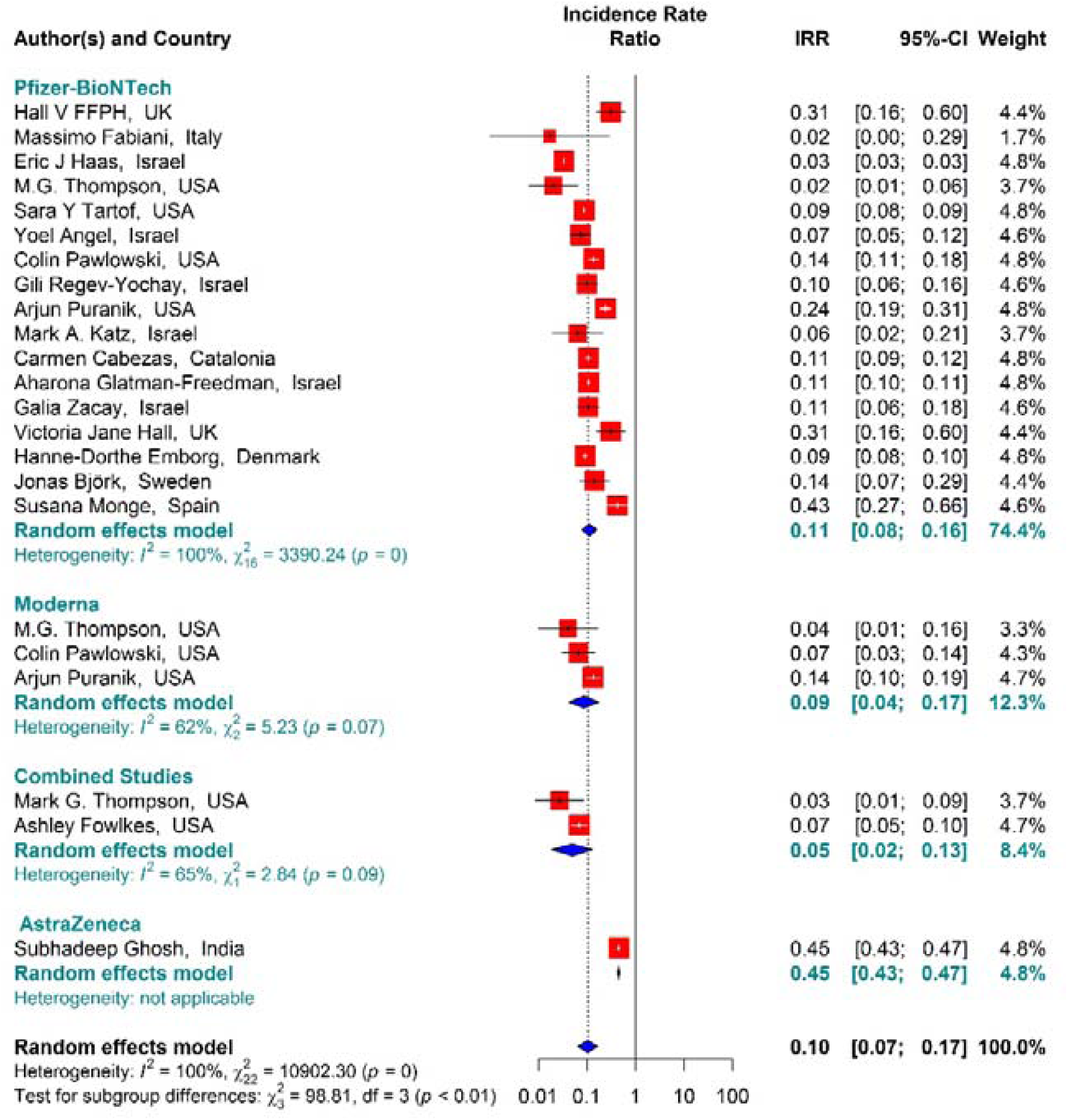
Effectiveness of vaccines using COVID-19-related IRR in full vaccinated individuals

### Effectiveness of vaccines against COVID-19 infection using Hazard ratio in partial-vaccinated individuals

Studying the Hazard ratio associated with SARS-COV 2 infection showed that vaccination with the first dose of BNT162b2 mRNA, mRNA-1273, ChAdOx1, and Combined studies reduced the risk of SARS-COV 2 infection by about 69% (HR = 0.31, 95% CI: 0.20-0.46) (Figure 7). In other words, the first doses of BNT162b2 mRNA, mRNA-1273, and ChAdOx1 vaccines had reduced the risk ratio of SARS-COV 2 infection by about 70% (HR = 0.30, 95% CI: 0.19-0.47), 83% (HR = 0.17, 95% CI: 0.05-0.59), and 39% (HR = 0.61, 95% CI: 0.51-0.72), respectively. On the other hand, the combined studies had reduced the risk of SARS-COV 2 infection by about 83% (HR = 0.17, 95% CI: 0.03-1.01). The results of the sub group analysis in the first dose showed well that there was a difference between the effectiveness rates of different types of vaccine against SARS-COV 2 infection (*p-value*_*subgroup*_ < 0.01) (Figure 7).

**Figure 7-.**
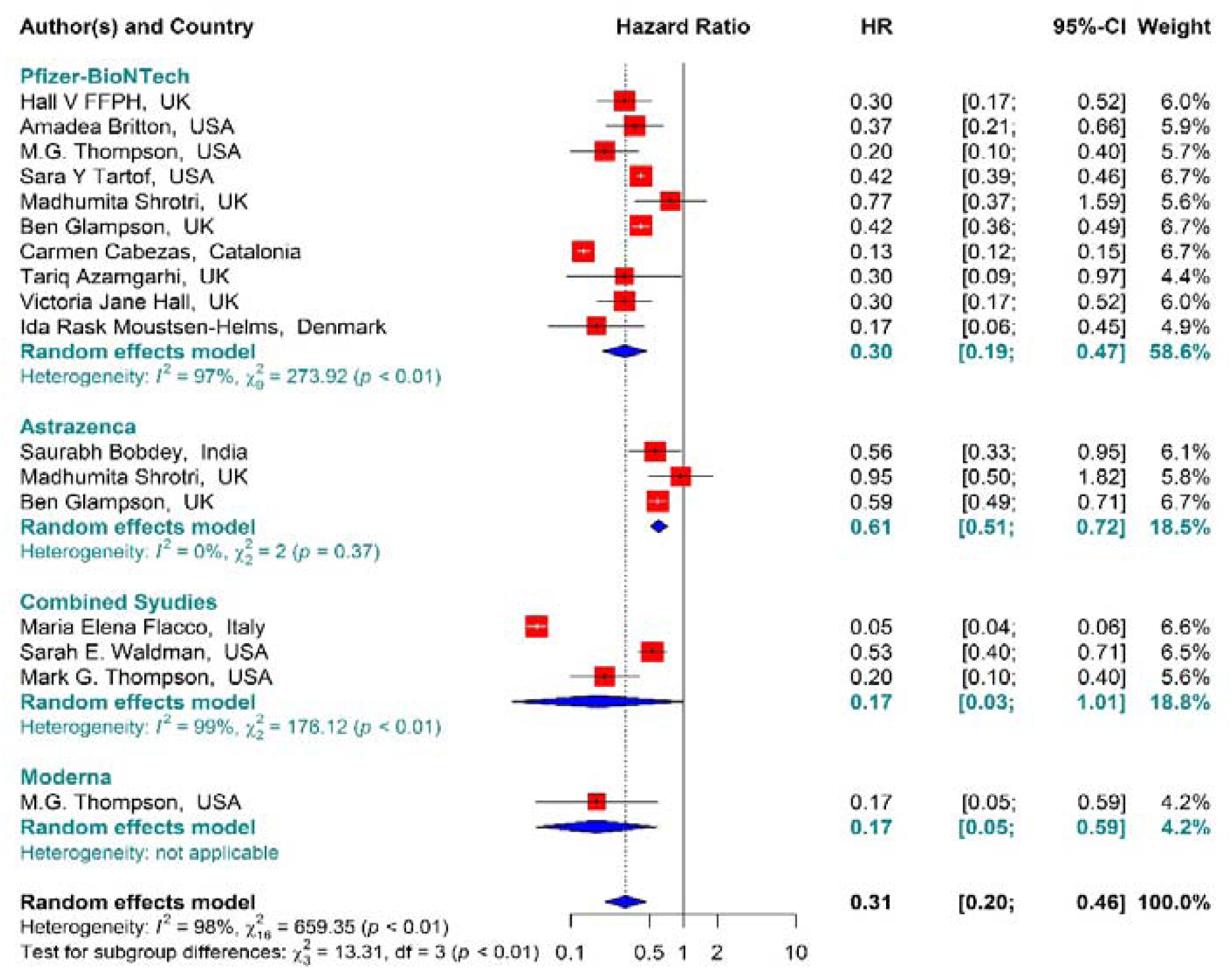
Effectiveness of vaccines against COVID-19 infection using Hazard ratio in partial-vaccinated individuals

### Effectiveness of vaccines against COVID-19 infection using Hazard ratio in full-vaccinated individuals

In the individuals vaccinated with the second dose (Full vaccinated) of BNT162b2 mRNA, mRNA-1273, and ChAdOx1 vaccines as well as the combined studies, the risk of SARS-COV 2 infection had reduced by about 84% (Figure 8). However, the second dose of BNT162b2 mRNA, mRNA-1273, and ChAdOx1 vaccines had reduced the Hazard ratio by about 79% (HR = 0.21, 95% CI: 0.14-0.31), 87% (HR = 0.13, 95% CI: 0.11-0.15), and 86% (HR = 0.14, 95% CI: 0.05-0.42), respectively. Furthermore, the Combined studies had reduced the risk of SARS-COV 2 infection in the individuals vaccinated with a second dose by about 90% (HR = 0.10, 95% CI: 0.03-0.34). The results of the sub group analysis in the second dose showed well that there was no difference between the effectiveness rates of different types of vaccines (*p* − *value*_*subgroup*_ = 0.2) (Figure 8).

**Figure 8-.**
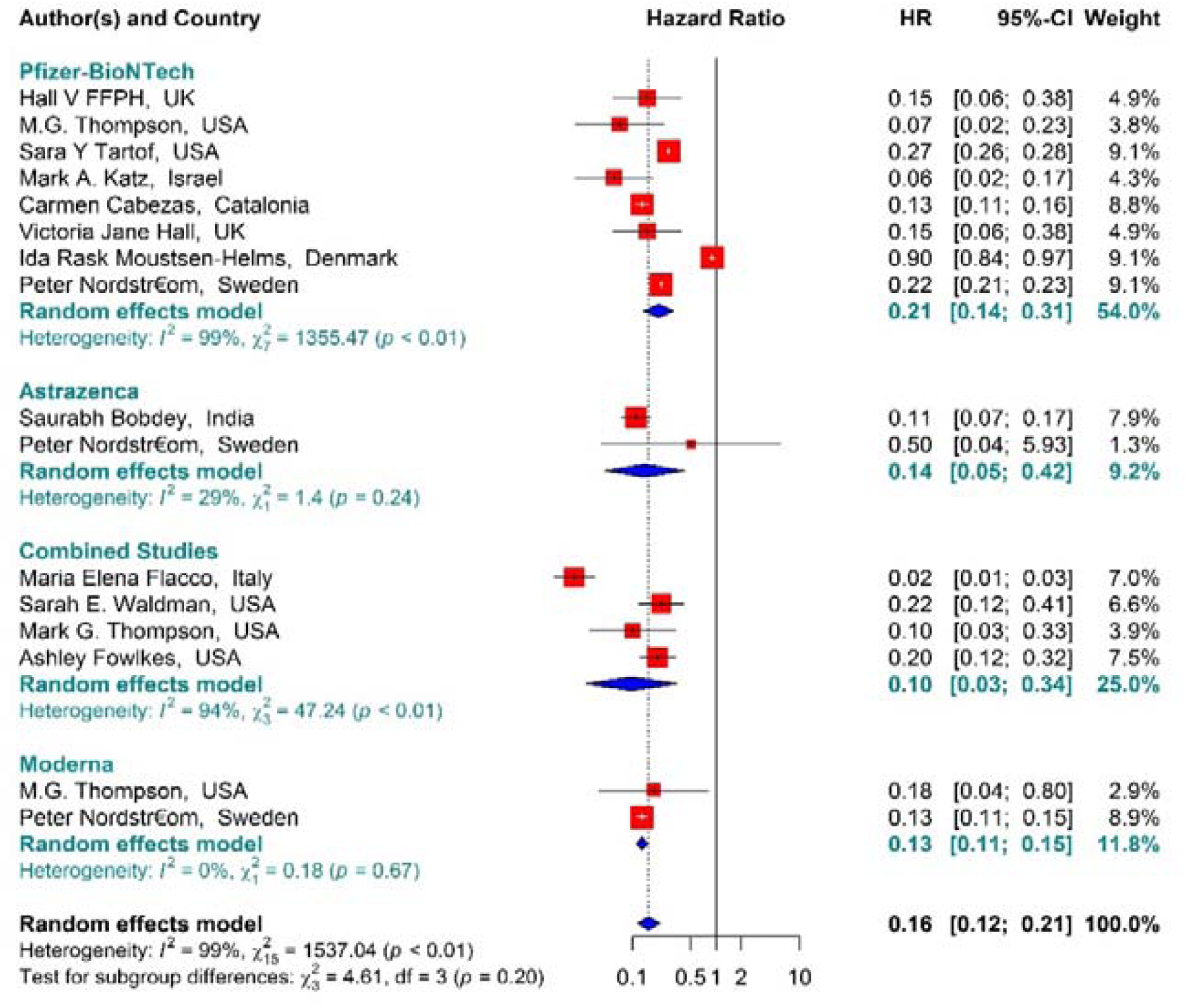
Effectiveness of vaccines against COVID-19 infection using Hazard ratio in full vaccinated individuals

### Effectiveness of vaccines against COVID-19-related mortality using Hazard ratio in partial-vaccinated individuals

The COVID-19-associated mortality Hazard ratio in the first-dose vaccinated individuals presented in Figure 13 showed well that the first-dose vaccination with BNT162b2 mRNA, ChAdOx1, and Combined studies had reduced the COVID-19-related mortality rate by about 68% (HR = 0.32, 95% CI: 0.23-0.45). However, the people vaccinated with the first dose of BNT162b2 and ChAdOx1 showed 58% (HR = 0.42, 95% CI: 0.30-0.59) and 61% (HR = 0.39, 95% CI: 0.23-0.68) reduction in the mortality Hazard ratio. Besides, the combined studies had reduced the risk of COVID-19-related mortality by about 91% (HR = 0.09, 95% CI: 0.01-0.64). The results of the sub group analysis in the first dose showed well that there was no difference between the effectiveness of different types of vaccines against COVID-19-related mortality rates (*p* − *value*_*subgroup*_ = 0.31) (Figure 9).

**Figure 9.**
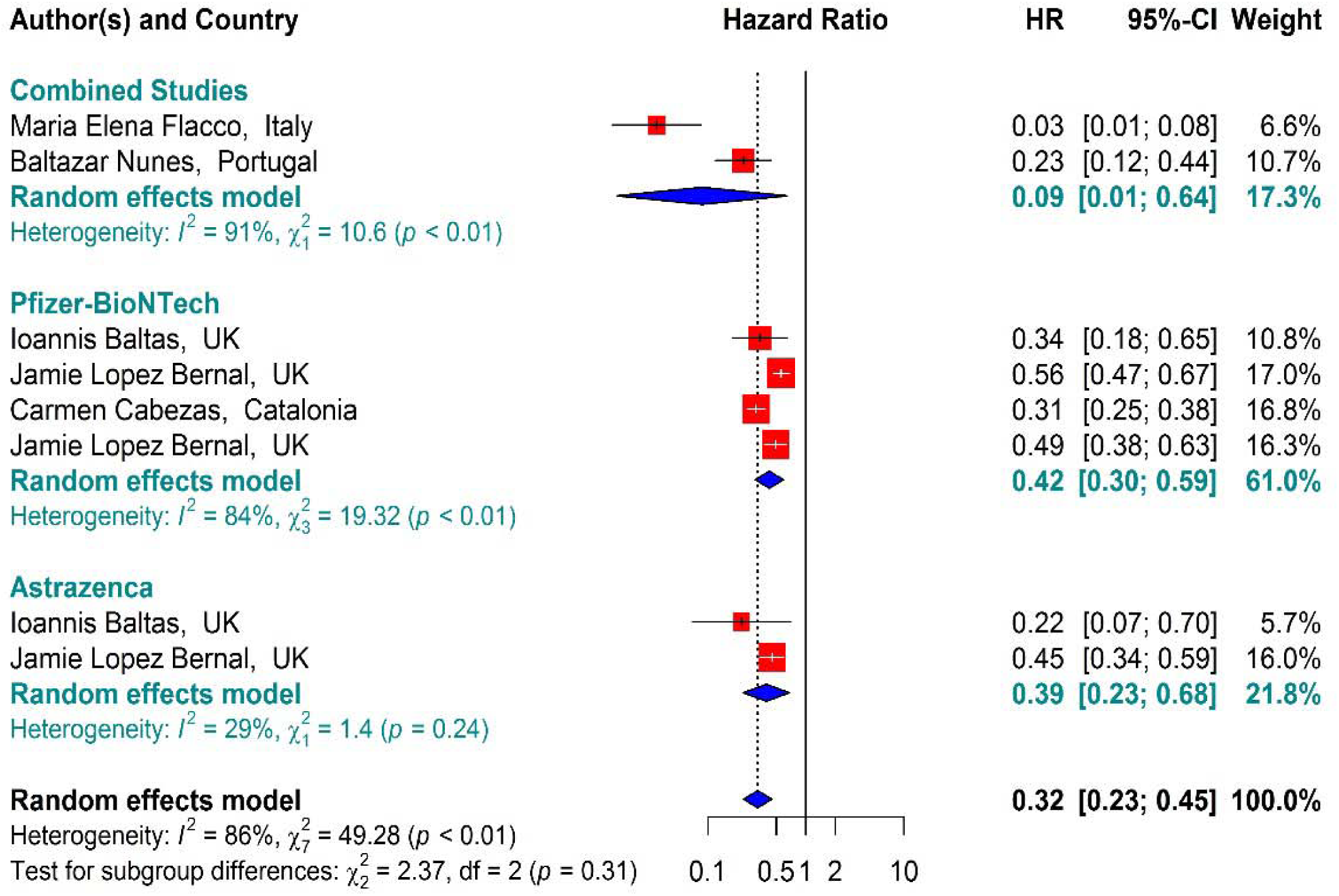
Effectiveness of vaccines against COVID-19-related mortality using Hazard ratio in partial-vaccinated individuals

### Effectiveness of vaccines against COVID-19-related mortality using Hazard ratio in full-vaccinated individuals

In the individuals fully vaccinated with BNT162b2 mRNA as well as combined studies, the COVID-19-associated mortality Hazard ratio was reduced by approximately 89% (HR = 0.11, 95% CI: 0.03-0.43). However, BNT162b2 mRNA vaccine and the Combined studies had reduced the risk by about 85% (HR = 0.15, 95% CI: 0.02-0.90) and 96% (HR = 0.04, 95% CI: 0.02-0.08), respectively. The results of the sub group analysis in the second dose showed that there was no difference between the effectiveness of different vaccines against COVID-19-related death (*p* − *value*_*subgroup*_ = 0.19) (Figure 10).

**Figure 10.**
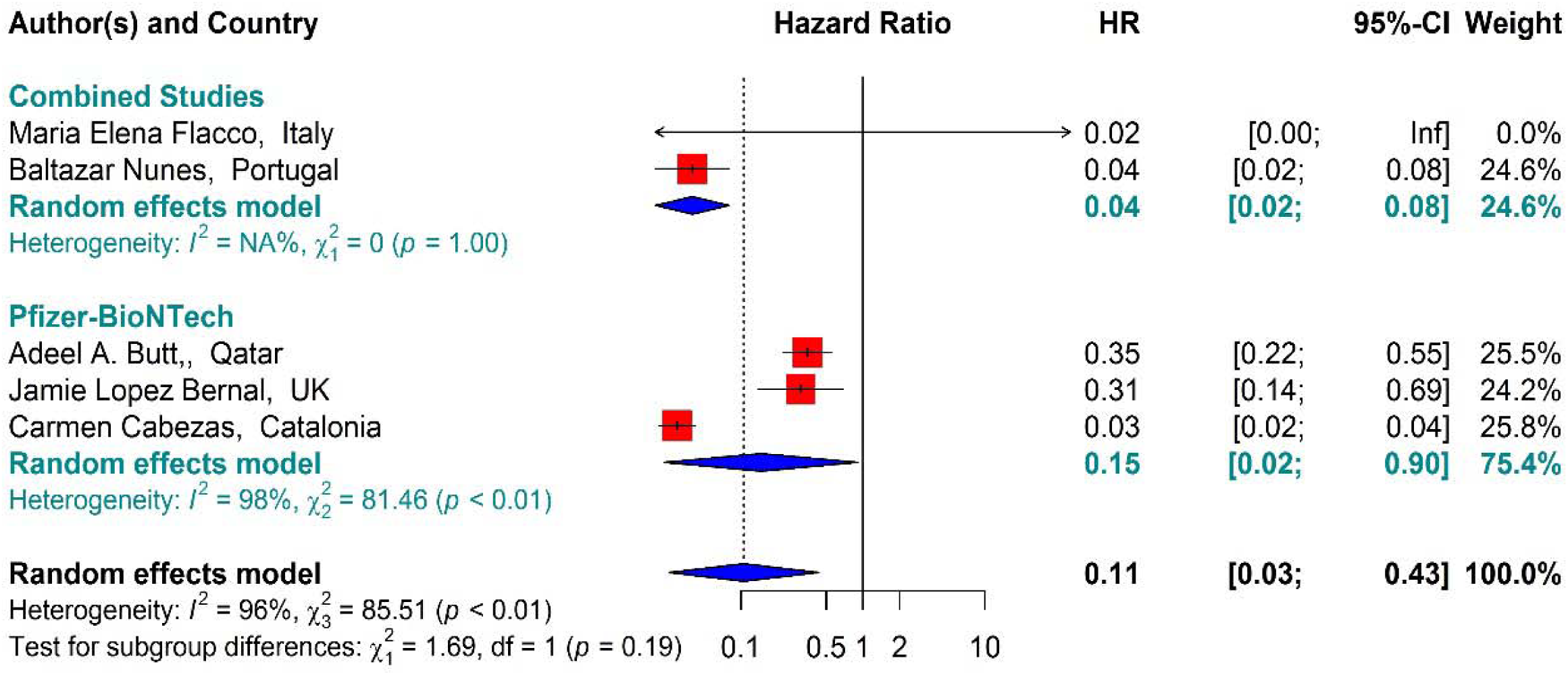
Effectiveness of vaccines against COVID-19-related death using Hazard ratio in full-vaccinated individuals

### Effectiveness of vaccines against COVID-19-related death using IRR of Partial- and Full-vaccinated individuals

The results of examining the effectiveness of the first dose of vaccines against COVID-19-related mortality showed that the mortality rate in the people vaccinated with the first dose of BNT162b2 mRNA, mRNA-1273, ChAdOx1, and Combined studies had reduced by about 25% (IRR = 0.75, 95% CI: 0.19-2.93) (Figure 11).

**Figure 11.**
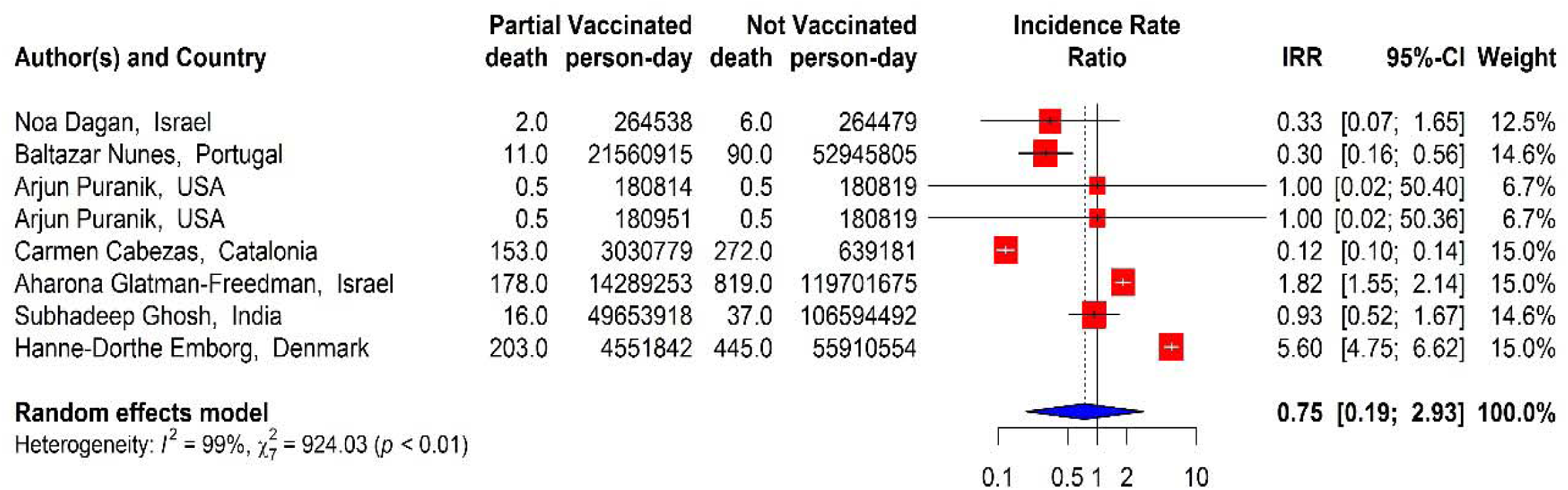
Overall effectiveness of vaccines against COVID-19-related mortality in first-dose vaccinated subjects

In addition, the effectiveness of BNT162b2 mRNA, mRNA-1273, and ChAdOx1 vaccines as well as the Combined studies against COVID-19-related mortality in the second dose was about 82% (IRR = 0.18, 95% CI: 0.15-0.34) (Figure 12).

**Figure 12.**
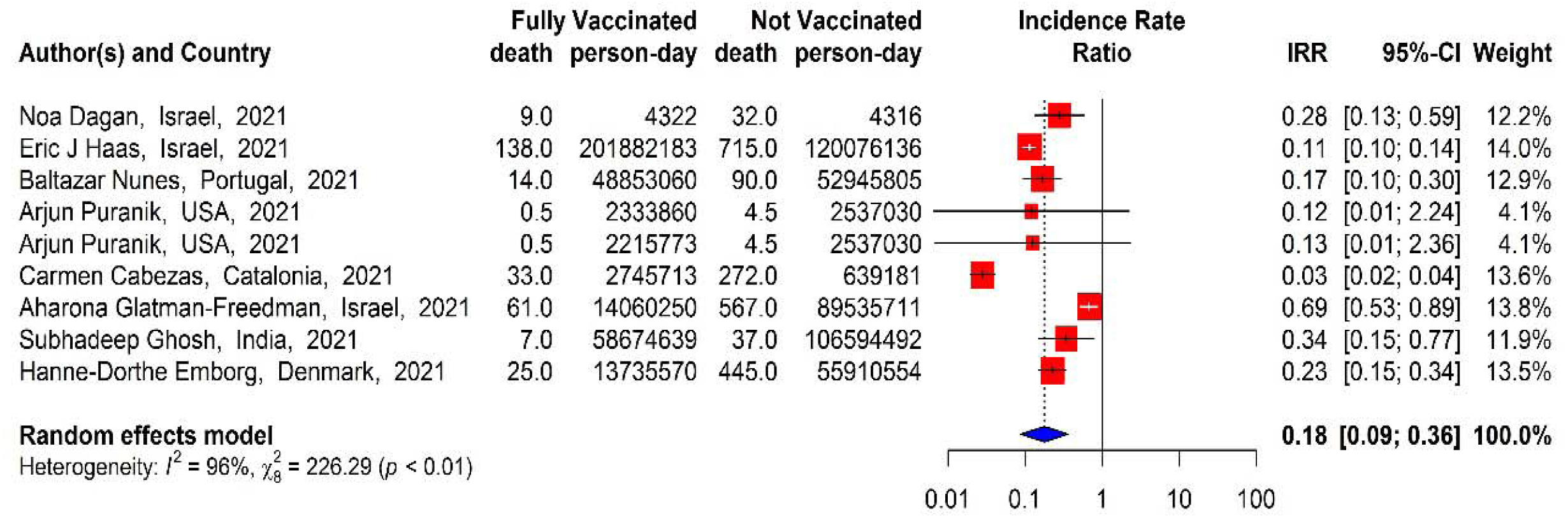
Overall effectiveness of vaccines against COVID-19-related mortality in second-dose vaccinated subjects

## Discussion

This meta-analysis, which aimed to evaluate the effectiveness of vaccination on the incidence of SARS-COV 2 infection as well as the post-vaccination mortality and hospitalization, was in line with the results of Phase 3 of the Clinical trials on the effectiveness of BNT162b2 mRNA, mRNA-1273, and ChAdOx1 vaccines (10, 21, 22). In other words, the effectiveness of the first and second doses of BNT162b2 mRNA vaccine against SARS-COV 2 infection was 82% and 95%, respectively (10), and in the present study, the effectiveness against SARS-COV 2 infection based on the Pooled estimate was 72% and 89%, respectively. The effectiveness of ChAdOx1 and mRNA-1273 vaccines against the incidence of infection was estimated at 51% and 69% in the first dose and 84% and 80% in the second dose, respectively, which is consistent with the previous studies (21-23).

The increased effectiveness of vaccination against SARS-COV 2 infection after the second dose was clearly observed in this study consistent with the results of other studies. In other words, the increased effectiveness of all vaccines against SARS-COV2 infection was generally about 16% (71% effectiveness in the first dose and 87% in the second). The increased effectiveness of BNT162b2 mRNA vaccine in the second dose compared to the first one was about 15%, and that of mRNA-1273 and ChAdOx1 vaccines was 11% and 33%, respectively. The difference between the effectiveness of two doses of vaccines against the incidence of SARS-COV2 infection in the studies that examined the vaccines heterogeneously (a combination of COVID-19 vaccines on the general population) was about 11% as well. Although the present study aimed to evaluate the effectiveness of homologous vaccines, there were some studies that examined a combination of populations vaccinated with BNT162b2 mRNA, mRNA-1273, and ChAdOx1, and even Ad26.COV2.S in very rare cases, the results of which showed significant effectiveness of the vaccines. That is to say, the effectiveness of these vaccines in reducing the incidence of infection, hospitalization, and mortality associated with COVID-19 in the vaccinated individuals was much higher than other vaccines in some doses. In a heterologous study of the vaccines, Nordstrom et al. (24) showed that their effectiveness varied 67% to 79% depending on their types. The results of the combined studies on the populations vaccinated with different types of vaccines were also consistent with the study by Nordstrom et al, and strengthened the hypothesis of the effect of heterologous vaccines against SARS-COV 2 infection.

On the other hand, the effectiveness of the vaccines increased significantly with the increased post-vaccination follow-up periods, especially after the second dose, so that Hunter et al. (25) reported high effectiveness of the first dose of BNT162b2 mRNA twenty-one days after injection. In the present study, 7≤ days after the first dose and ≥ 5 days after the second dose were considered as partial and full vaccinated, respectively. The results indicated the high effectiveness of the second dose of COVID-19 vaccines (which varied between 20 and 30 days after the first dose) against COVID-19 infection, hospitalization, and mortality.

Although the effectiveness of COVID-19 vaccines against alpha and delta variants has been reported to be lower and milder, the effectiveness of full vaccination against these variants has been shown to be high in other studies (26). In a similar study, Haas et al. (17) reported high effectiveness of two doses of BNT162b2 mRNA vaccine against SARS-COV2 infection, hospitalization, and mortality in B.1.1.7 variant. In addition, some studies on the effects of COVID-19 vaccines on Delta variant did not observe a significant effect 28 days after the first dose (27). This study, which examined the effectiveness of COVID-19 vaccines regardless of SARS-COV2 variants, also showed the effect of complete vaccination on reducing the incidence of infection, hospitalization and mortality.

The effectiveness of complete COVID-19 vaccination in reducing the rate of hospitalization in this study was well demonstrated, consistent with the results of other studies on prevention of COVID-19-related hospitalization (17, 19, 28). The effectiveness against COVID-19-associated hospitalization and mortality was significantly increased with the second dose according to the immunological responses of the subjects, such that the biggest difference in the effectiveness of the two vaccine doses against hospitalization was related to BNT162b2 mRNA and mRNA-1273 with 35% and 29% increase in the second doses, respectively. Reduced mortality rates was another aspect of the effectiveness of COVID-19 vaccines, i.e. the second dose injection was associated with a further 21% decrease in the risk of COVID-19 mortality compared to the first dose (68% in the first dose vs. 89% in the second).

Despite the important results found out on the effectiveness of COVID-19 vaccines against the incidence of infection, hospitalization, and mortality after vaccination, the present study had a number of limitations, including the possibility of vaccination of specific age or occupational groups (health care worker) associated with an increase or decrease in the chance of contracting SARS-COV2 infection. However, the confounding effect of the background variables was somewhat controlled through the use of HR adjusted studies. Herd immunity was another issue that could distort the overall results; i.e. with increasing follow-ups, the risk of infection in the community and non-vaccinated people would gradually decrease, and this would overestimate the effectiveness of various COVID-19 vaccines.

## Conclusion

The results of this study showed that vaccination with BNT162b2 mRNA, mRNA-1273, and ChAdOx1, and even their combination in different populations was associated with increased effectiveness against SARS-COV2 infection, hospitalization, and mortality in the first and second doses. On the other hand, due to the higher effectiveness of the second dose compared to the first one in reducing the incidence of infection, mortality, and hospitalization associated with COVID-19, injection of the second and the booster doses seems necessary for high risk individuals. Furthermore, considering the increasing trend of vaccine effectiveness, the use of other preventive methods such as social distancing and wearing facemasks, preferably 0-7 days after the first dose, seems necessary.

## Data Availability

All data produced in the present study are available upon reasonable request to the authors

